# Accelerating vaccine trials during an outbreak of Disease-X: the effect of pathogen super-spreading on ring-trial design

**DOI:** 10.64898/2026.02.17.26346480

**Authors:** Robert Hinch, Ian Roberts, Chris Wymant, Lucie Abeler-Dörner, Sarah Lapidus, Marc Lipsitch, Christophe Fraser

## Abstract

The prospective design of vaccine efficacy trials for deployment in outbreaks requires advance consideration of plausible outbreak scenarios, anticipated vaccine characteristics, and logistical and ethical constraints. As part of CEPI’s 100 Days Mission to accelerate vaccine development against a novel Disease X, we evaluated trial designs for a hypothetical Nipah-X outbreak. We assumed Nipah-X would share key features with Nipah, including high case fatality rates and substantial super-spreading, but with sustained human-to-human transmission. Using simulations based on infection models, including an extended chain-binomial model incorporating super-spreading, we compared ring-trials using cluster-randomisation with individual-randomisation within rings. High levels of super-spreading markedly reduced the power of cluster-randomised designs due to strong intra-cluster correlations in case numbers, whereas individual-randomisation retained power. These findings highlight that understanding and accounting for super-spreading is critical when designing ring-trials, as cluster-randomised designs may fail unless vaccine efficacy is nearly complete.

## INTRODUCTION

The CEPI 100 Days Mission aims to develop and license vaccines against novel pathogens with pandemic potential within 100 days of pathogen identification (CEPI, 2022). A central component of this initiative is the development of vaccine candidates against exemplar viruses from diverse families (Gouglas et al., 2019) using adaptable platform technologies such as mRNA (Pardi et al., 2018) and ChAdOx (Antrobus et al., 2014). While these platforms can generate vaccine candidates within weeks, demonstrating efficacy through field trials can take many months (Baden et al., 2020; Voysey et al., 2021) or even years (Datoo et al., 2024). Selecting an optimal trial design is therefore critical to accelerate licensure of a vaccine with proven efficacy.

The choice of vaccine efficacy trial design is largely determined by the epidemiology of the pathogen (Dean et al., 2019). For pathogens that reach high prevalence in the general population, such as SARS-CoV-2, population-based individually-randomised controlled trials remain the gold standard. In contrast, pathogens with very high infection fatality rates typically exhibit low prevalence because strict public health measures limit transmission, as seen with Ebola virus, and therefore targeted designs are required to accrue sufficient cases within the trial cohort (Dean et al., 2019).

Nipah virus is a highly virulent pathogen that causes small annual outbreaks in Bangladesh and southern India (Yadav et al., 2025). Most human infections result from consumption of date-palm sap contaminated by bat excreta, with limited or no human-to-human transmission (Nikolay et al., 2019). However, occasional large super-spreading events involving human-to-human transmission have been documented (Arunkumar et al., 2020). Several Nipah vaccine candidates have been developed (DeBuysscher et al., 2016; van Doremalen et al., 2022) and are now undergoing Phase I safety and immunogenicity trials (Rodrigue et al., 2024).

Although current outbreak sizes are too small to support an efficacy trial (Nikolay et al., 2021), there is concern that the virus could evolve increased human-to-human transmissibility, resulting in larger outbreaks. We refer to such a hypothetical variant as Nipah-X. As part of CEPI’s 100 Days Mission, in collaboration with PATH (CEPI, 2023), a protocol for a potential Nipah-X vaccine efficacy trial is being developed. The work presented here informed this protocol. In our analyses, Nipah-X was assumed to retain the high virulence and substantial super-spreading observed in Nipah virus, while outbreaks would remain self-limiting under public-health interventions, with total sizes between approximately 3,000 and 10,000 cases.

A natural starting point for designing a vaccine efficacy trial is to consider previous trials of pathogens with similar epidemiological characteristics. Given the assumed high virulence and limited outbreak size of Nipah-X, its epidemiology would likely resemble that of Ebola virus during the 2014–2016 West African outbreak, which resulted in approximately 28,600 infections and 11,325 deaths (Jacob et al., 2020). During this outbreak, the *Ebola Ça Suffit!* trial successfully demonstrated the efficacy of an rVSV-vectored vaccine using a cluster-randomised ring-trial design (Henao-Restrepo et al., 2017). In a ring-trial, participants are recruited based on their contact with a known index case, typically identified through contact tracing, under the rationale that infection risk is substantially higher among contacts than in the general population (Butzin-Dozier et al., 2022). Although this design increases case accrual, some participants may already be infected at enrolment and develop symptoms shortly after randomisation. Vaccines are generally less effective if administered post-infection, therefore cases arising within a defined “exclusion window” after randomisation are typically omitted from efficacy analyses. In the *Ebola Ça Suffit!* trial, this window was set at 10 days: vaccine efficacy was negligible within this period but reached an estimated 100% afterwards (Henao-Restrepo et al., 2017). The trial used cluster-randomisation, assigning each ring either to immediate vaccination or to a delayed (21-day) arm, and its success has led cluster-randomised ring-trials to be the default WHO protocol for filoviruses (WHO, 2024). An alternative trial design is to individually randomise participants within each ring to the immediate or delayed vaccination (or placebo) arm. Assessing the relative merits of these randomisation strategies, and the epidemiological conditions under which their performance diverges, is the focus of this study.

Here, we model vaccine ring-trials for pathogens exhibiting varying degrees of super-spreading and vaccine efficacy. A key determinant of the power of cluster-randomised trials is the intra-cluster correlation (ICC; Killip et al., 2004). We show that pathogens characterised by high levels of super-spreading generate correspondingly high ICCs, which severely reduce statistical power unless vaccine efficacy is nearly complete. Beyond the direct protection conferred to vaccinated individuals, vaccines that block transmission can also provide indirect protection by lowering overall infection rates within highly vaccinated groups (i.e., herd immunity). Under such circumstances, vaccine *effectiveness* in reducing infections is higher than the vaccine *efficacy* of protecting an isolated individual. Cluster-randomised designs, which measure this higher combined vaccine *effectiveness*, gain additional power, whereas individual-randomised designs measure only direct *efficacy* (Longini et al., 1993). However, we demonstrate that while this indirect effect improves the performance of cluster-randomised trials for pathogens without pronounced super-spreading, its impact is small compared with that of super-spreading.

## RESULTS

Pathogens vary widely in the extent of super-spreading observed during transmission, and licensed vaccines against them span a broad range of efficacies (Figure 1). Super-spreading reflects the degree of dispersion in the offspring distribution of secondary cases and is typically quantified by fitting a negative-binomial model to transmission data (Lloyd-Smith et al., 2005). The dispersion parameter *k* captures this variability: small values of *k* indicate highly overdispersed offspring distributions and substantial super-spreading. For example, *k = 0.1* corresponds to a situation in which roughly 20% of infected individuals account for 80% secondary cases.

**Figure 1.**
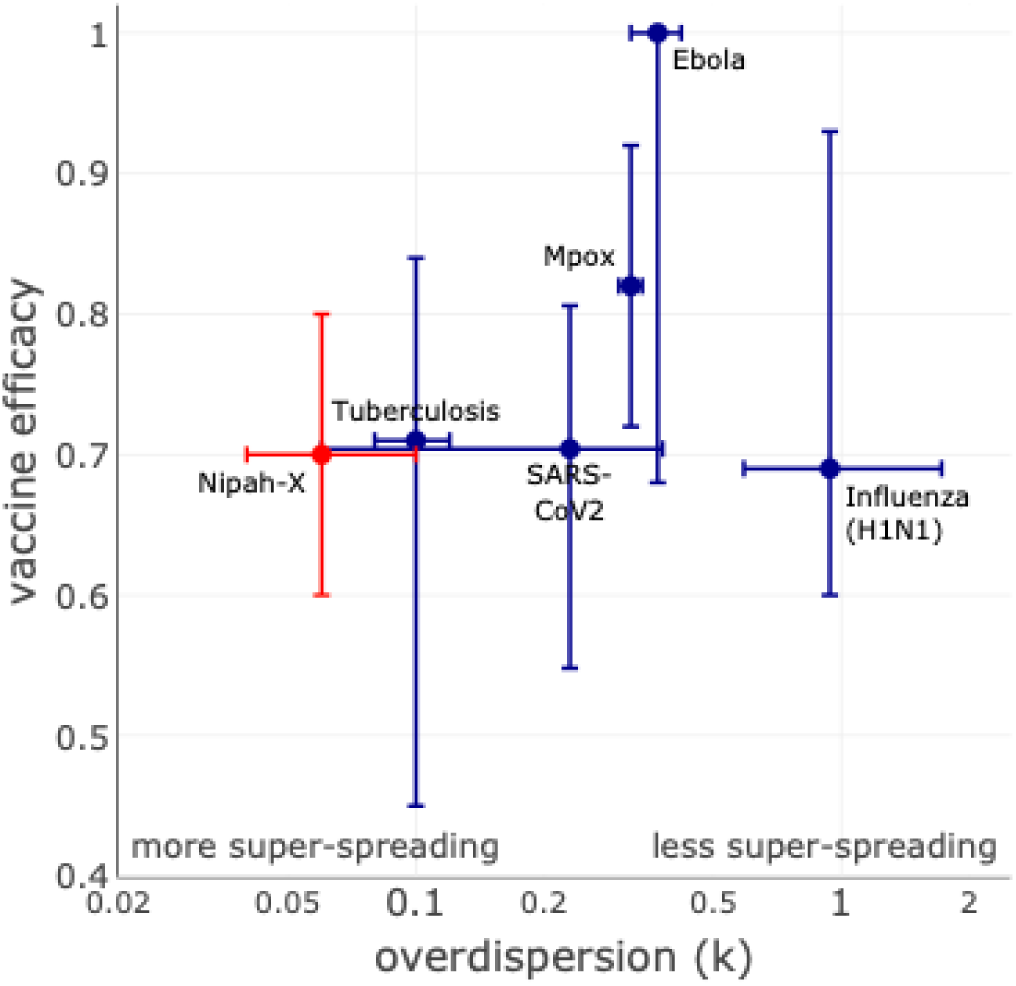
Vaccine efficacy and super-spreading across infectious pathogens. Vaccine efficacy is shown against the overdispersion parameter (*k*), estimated from negative-binomial fits to offspring distributions describing transmission heterogeneity. Most pathogens exhibit substantial super-spreading (*k* < 0.5), with Ebola showing moderate overdispersion (*k* ≈ 0.37) but unusually high vaccine efficacy. The figure also includes our baseline assumption for Nipah-X, based on reported *k* estimates for Nipah virus. Estimates of *k* are taken from Ebola (Lau et al., 2017), mpox (Maniscalco et al., 2024), SARS-CoV-2 (Hodcroft et al., 2025), tuberculosis (Ypma et al., 2013), H1N1 influenza (Fraser et al., 2011), and Nipah virus (Nikolay et al., 2021). Vaccine efficacy estimates are from Ebola (Henao-Restrepo et al., 2017), mpox (Piscel et al., 2024), SARS-CoV-2 (Voysey et al., 2021), tuberculosis (Colditz et al., 1994), and H1N1 influenza (Osterholm et al., 2012).

Figure 1 summarises the reported levels of super-spreading and corresponding vaccine efficacies for several pathogens. Our baseline assumption for Nipah-X is that it exhibits levels of super-spreading similar to those of Nipah virus, and that vaccine efficacy is set to a standard value commonly used in trial power calculations. Although Nipah is associated with high levels of super-spreading, it is not an outlier; elevated super-spreading (*k < 1*) is common among many pathogens. These empirical patterns illustrate the range of transmission heterogeneity observed across pathogens. To understand how such variability influences the feasibility and design of vaccine efficacy trials, we developed a simulation framework to explore the performance of alternative ring-trial designs under differing levels of super-spreading and vaccine efficacy.

### Super-Spreading Induces Intra-Cluster Correlation

The intra-cluster correlation (ICC) measures the similarity of outcomes within a cluster relative to between clusters. High ICC values reduce the effective sample size and thereby lower the power of a cluster-randomised trial to detect vaccine efficacy (Killip et al., 2004). For pathogens exhibiting super-spreading, a small proportion of index cases generate a large share of infections, leading to the concentration of cases in a few rings (Figure 3A) and consequently a high ICC.

To quantify this effect, we modelled individual rings using the chain-gamma-binomial model (Fraser et al., 2011). In this model, each ring is treated as an isolated community with homogeneous internal mixing (Figure 2), and the infectiousness of each infected individual follows a gamma distribution. Rings were assumed to contain 30 individuals and ICCs were calculated (Chakrobarty et al., 2018) across a range of super-spreading levels and ring attack rates (AR). As shown in Figure 3B–C, ICC increased with both super-spreading and AR.

**Figure 2:**
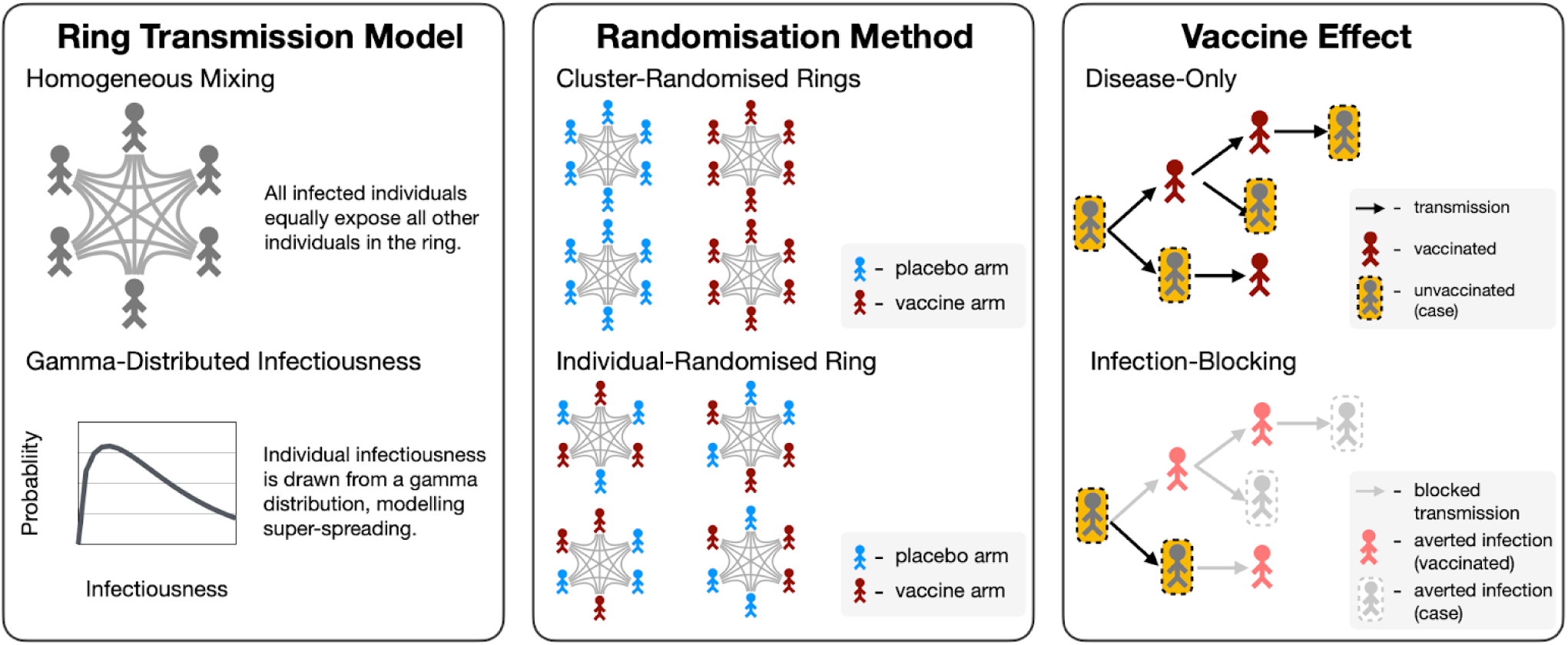
Model definition. The ring transmission model is the chain-gamma-binomial model, which assumes homogeneous mixing within a ring and gamma-distributed individual infectiousness. With cluster-randomisation, all individuals in a single ring are either in the vaccine or placebo arm. With individual-randomisation, individuals within each ring can be in either the vaccine or placebo arm. *Disease-only* vaccines only prevent disease in vaccinated individuals, whereas the *infection-blocking* vaccines prevent onward transmissions thus averting infections in both vaccinated and unvaccinated individuals.

Using the estimated level of super-spreading for Nipah virus (Nikolay et al., 2021) and an AR of 3.33%, the predicted ICC was 35%, suggesting that super-spreading would substantially reduce the power of a cluster-randomised ring-trial. For the Ebola virus, which exhibits more moderate super-spreading (Lau et al., 2017), we simulated the delayed arms of the *Ebola Ça Suffit!* Trial (AR of 1.2%; 47 rings and 65 people per ring) which yielded a median ICC of 5.5% (95% PI:1%-18%). Although the central estimate is lower than the reported 14%, ICC estimates derived from small numbers of clusters are associated with high statistical uncertainty (Figure 3D), and the *Ebola Ça Suffit!* value lies within the 95% prediction interval of 1%-18%.

**Figure 3:**
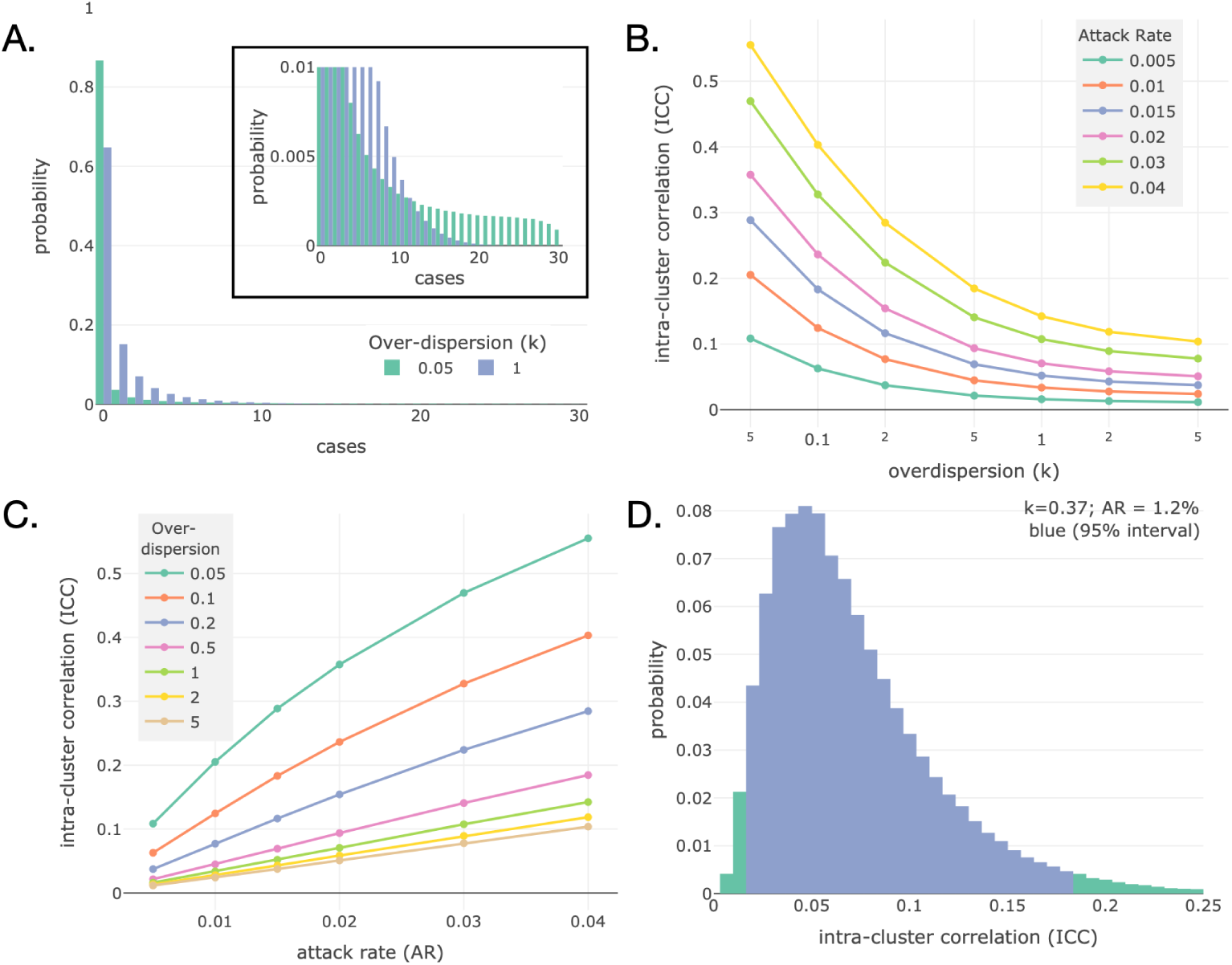
Super-spreading induces intra-cluster correlation. A. The distribution of cases in a ring of 30 individuals and a 3.33% AR, but different levels of super-spreading. With large amounts of super-spreading, the probability of zero cases is much higher, but the tail of distribution is much longer. The inset panel zooms in on the main plot. B. The ICC for clusters with different levels of super-spreading (log-scale axis) and different AR. The ICC increases dramatically as k is reduced below 0.2. C. The ICC for clusters with different AR (linear axis) and different levels of super-spreading. The ICC increases with the AR. D. The distribution of the maximum-likelihood estimates of ICC for simulated trials of 47 rings each containing 65 individuals, using the AR from the *Ebola Ça Suffit!* trial and the estimate of k for Ebola. The 95% central interval spans 1%-18% demonstrating the large variation in ICC which can be observed in trials.

Most trial designs include an exclusion period immediately after randomisation to omit individuals likely infected before vaccination (Henao-Restrepo et al., 2017). To account for this, we extended the chain-gamma-binomial model to exclude infections directly caused by the index case from the final size distribution. Repeating the calculations produced nearly identical relationships between ICC, super-spreading, and AR, despite a 65% reduction in total cases (Figure S1). When cases within 10 days of randomisation were excluded, the *Ebola Ça Suffit!* trial reported an ICC of 3.5%, considerably lower than the 14% estimate including all cases. We simulated the delayed arms of the *Ebola Ça Suffit!* Trial excluding the first-generation, which yielded a median ICC of 5.0% with a 95% prediction interval of 0%-20% containing the observed ICC in the trial. Note the very high statistical uncertainty inherent in ICC estimates derived from small numbers of clusters.

### Estimating Power of Ring-Trials in Response to Outbreaks

To evaluate the power of ring-trial designs, we assessed whether simulated trials estimated a vaccine efficacy of at least 30% with a Type I error ≤ 5%. In conventional individually randomised efficacy trials, where participants are drawn from the general population, power is typically expressed as the number of in-trial cases required to keep Type II error below a threshold (usually 20%). Although this remains an important benchmark for ring-trials, several additional factors must be considered.

First, cases in a ring-trial are expected to occur only within a short period following identification of the index case, particularly when placebo rings are vaccinated after a delay (Henao-Restrepo et al., 2017). This contrasts with conventional trials, in which participants are followed until enough cases accrue from general community transmission. Second, if vaccination prevents infection and transmission, indirect protection among contacts will reduce the total number of observed cases. This indirect effect, absent in standard iRCTs, depends on the randomisation procedure and must be accounted for when evaluating ring-trial designs. Finally, the number of rings that can be enrolled is often limited by logistical constraints, such as vaccine availability or outbreak control through other interventions, so trials may end before reaching the number of cases required by formal power calculations.

To capture these dynamics, we considered four complementary metrics of trial power: (1) the number of cases required to achieve 80% power; (2) the number of rings needed to reach that case threshold with 80% probability; (3) the probability of meeting the required number of cases for a fixed-size trial; and (4) the probability that vaccine efficacy is demonstrated in a fixed-size trial. Each metric was estimated across a range of vaccine efficacies and levels of super-spreading, assuming a constant mean attack rate in placebo rings. As a first step, we explored how super-spreading and vaccine efficacy together shape the power of cluster-randomised ring-trials, providing a baseline for comparison with alternative designs.

### Super-Spreading Reduces Power of Cluster-Randomised Design

We simulated cluster-randomised ring-trials using the chain-gamma-binomial model across a range of super-spreading levels and vaccine efficacies. As some vaccines can reduce transmission in addition to preventing disease, we modelled two vaccine types to capture the range of possible indirect effects: a *disease-only vaccine*, which prevents symptomatic disease without affecting transmission, and an *infection-blocking vaccine*, which completely prevents infection and onward spread (Fig 2). Most vaccines fall between these extremes; comparing them allows the contribution of indirect protection to be assessed relative to that of super-spreading. The vaccines were assumed to be *leaky*, *i.e.* reduce the probability of disease given infection (or infection given exposure) equally in all individuals, as opposed to *all-or-nothing*, *i.e.* some individuals have complete protection and some none. With low infection prevalence, the effect of *leaky* and a*ll-or-nothing* vaccines at the population level are almost equivalent.

Each simulated trial was statistically analysed using the Bayesian beta-binomial model, a frailty model in which the attack rate for each cluster is drawn from a beta distribution, and using the matched Bayesian chain-gamma-binomial model (see Methods). The beta-binomial model estimates the reduction in attack rate in the vaccine arm relative to the placebo arm; therefore, it measures vaccine *effectiveness* in cluster-randomised designs, because only the vaccine arm benefits from any indirect effect of vaccination. Conversely, in individual-randomised designs, the beta-binomial model measures vaccine *efficacy*, as participants in both arms benefit equally from the indirect effect of vaccination. For both cluster-randomised and individual-randomised designs, the matched Bayesian chain-gamma-binomial model estimates vaccine *efficacy* which is a model parameter, with indirect vaccine effects incorporated explicitly within the model. When using the matched Bayesian chain-gamma-binomial model, we therefore are making an *a priori* assumption about whether the vaccine blocks transmission.

As shown in Figure 4, power estimates from the four metrics were highly consistent for cluster-randomised ring-trials with *infection-blocking* vaccines. When super-spreading was limited (*k > 1*), trials were well powered if true vaccine efficacy exceeded ≈ 75%. In contrast, under high super-spreading (*k = 0.05*), trials were under-powered even when efficacy reached 95%. This reduction in power was consistent across all metrics and driven by the higher number of cases required as increasing super-spreading elevated the intra-cluster correlation (ICC). Because these trends were uniform, subsequent figures show only the probability of success for a fixed-size trial (200 rings of 30 individuals, AR = 3.33%), with the remaining metrics provided in the Supplementary Information.

**Figure 4:**
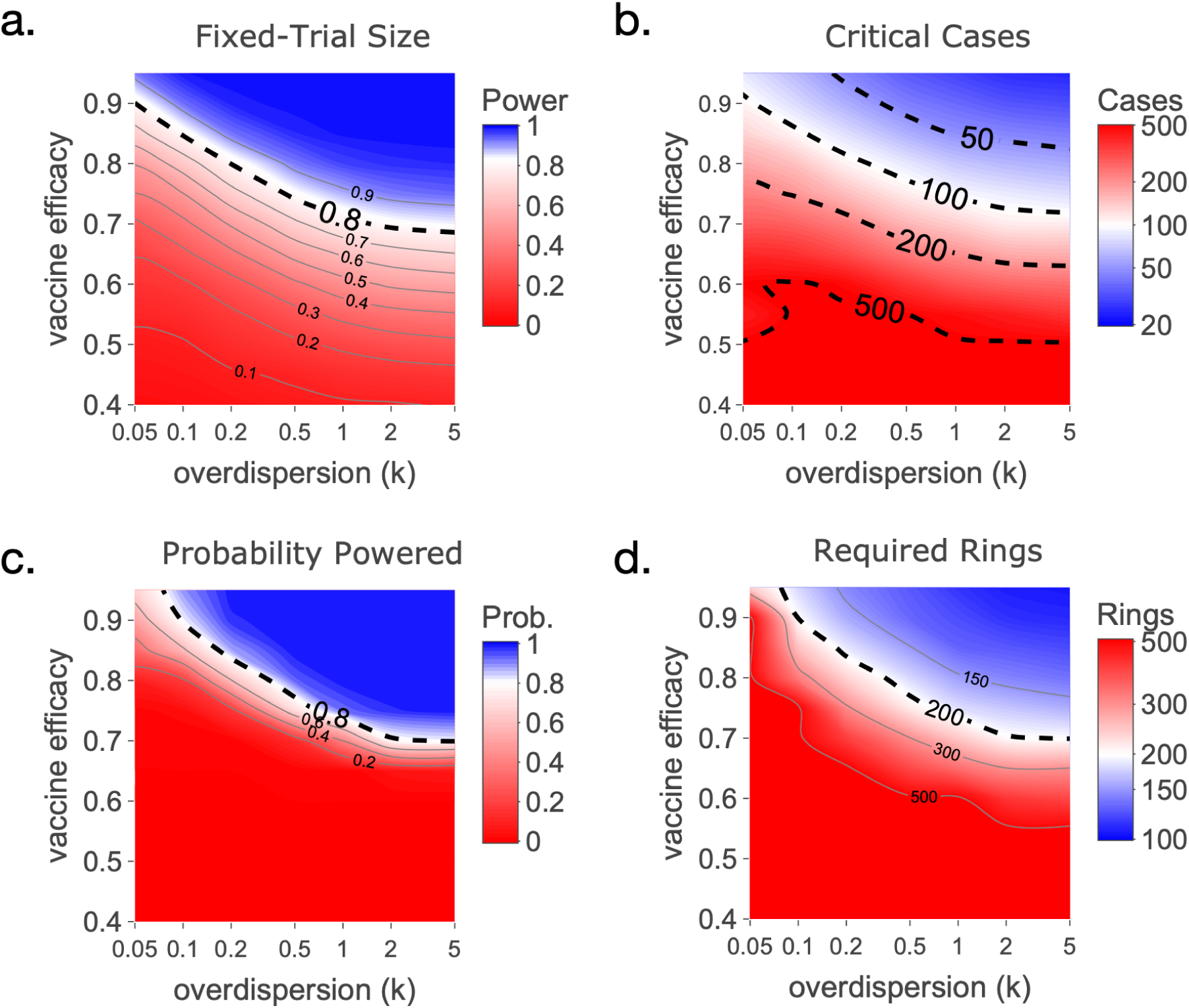
Measures of trial power. Due to how cases are accumulated in ring-trials, we consider different measures of trial power. a. the probability that a trial with a fixed number of rings (200 rings; 30 people per ring; AR 3.33%) is powered. b. the required number of cases across all rings such that the trial is 80% powered. c. the probability that a trial with a fixed number of rings accumulates the required cases. d. the required number of rings in the trial such that there is a 80% probability of accumulating the required cases. Each plot shows the trial power for different levels of super-spreading (lower k means higher super-spreading) and vaccine efficacies, with the blue representing the region where trial is powered.

Figure 5 compares the power of cluster-randomised designs for *disease-only* (top left panel) and *infection-blocking* vaccines (top right panel; all 4 metrics of power shown in Figure S4). At low super-spreading levels (*k > 1*), an *infection-blocking* vaccine achieved sufficient power at ≈ 55% efficacy, compared with ≈ 75% for a *disease-only* vaccine. This difference reflects the added indirect protection conferred by *infection-blocking* vaccines leading to the vaccine *effectiveness,* which is estimated by the beta-binomial model, being higher than the vaccine *efficacy*. However, as super-spreading intensified (*k < 0.2*), cluster-randomised designs became under-powered even for vaccines with high efficacy. The trial power was also calculated using a Bayesian analysis of the chain-gamma-binomial models (see Methods), which measures vaccine *efficacy*, where the action of the vaccine (e.g. *disease-only* or *infection-blocking*) is part of the model. For *disease-only* vaccines, where the vaccine *effectiveness* is the same as the vaccine *efficacy*, the two models give almost identical results (Figure S2, left column). For *infection-blocking* vaccines, the beta-binomial model appears slightly more powerful, which is due to it measuring the higher vaccine effectiveness (Figure S2, right column).

**Figure 5:**
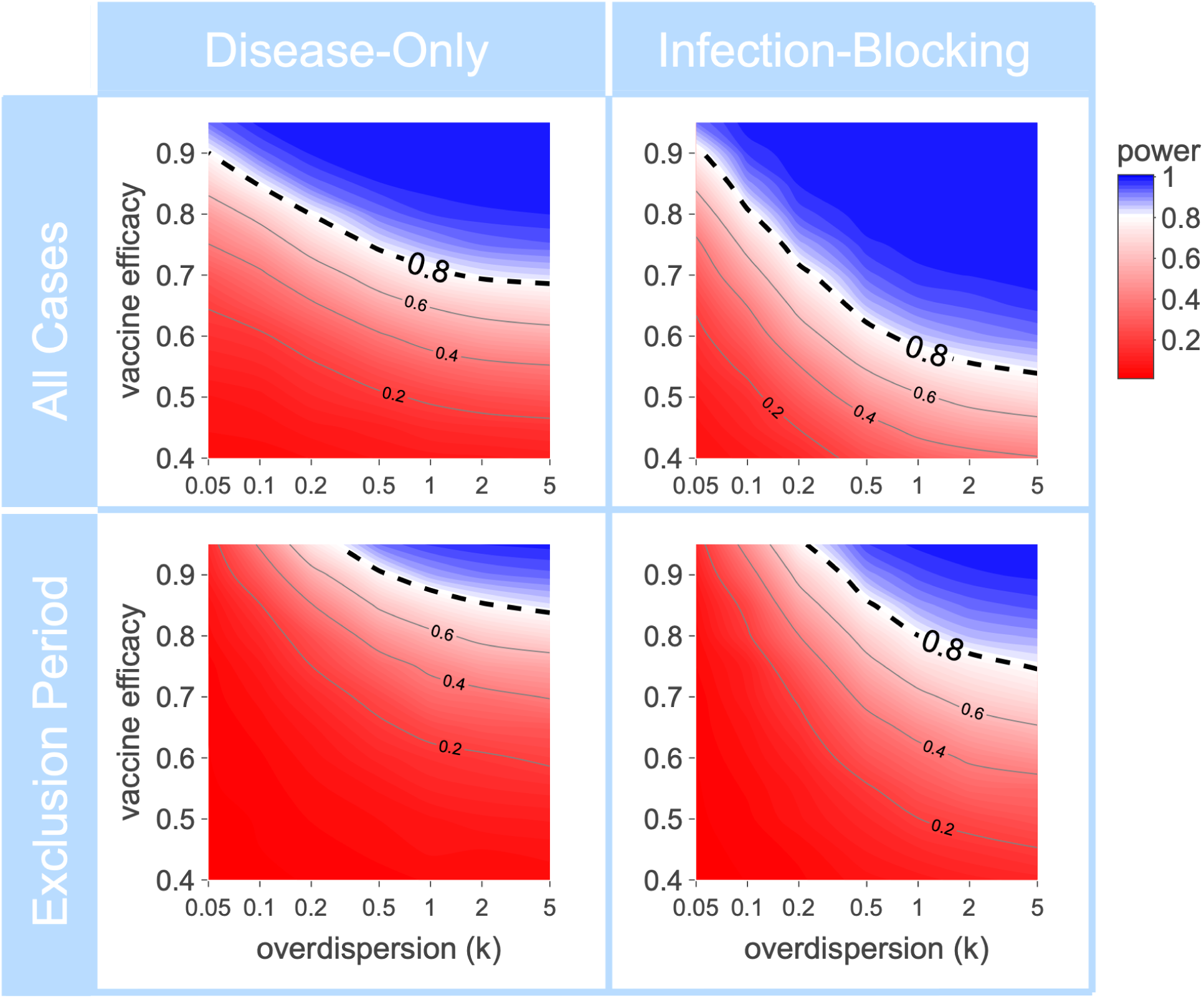
Super-spreading dramatically reduces the power of cluster-randomised trials. The probability of a fixed-size trial (200 rings; 30 per ring; 3.33% AR) demonstrating vaccine efficacy for different levels of super-spreading and vaccine efficacy. With high levels of super-spreading (k<0.1), the trial is always under-powered even if the vaccine efficacy is over 90%. The power of the trial is increased if the vaccine is *infection-blocking* (right column) as opposed to a *disease-only* vaccine (left column). The top row assumes the vaccine acts instantly and includes all cases. The bottom row assumes the vaccine is not instantly effective, and excludes cases who are infected directed by the index case.

Because vaccine trials typically include an exclusion window immediately after randomisation to omit participants likely infected beforehand (e.g. 10 days in the *Ebola Ça Suffit!* trial, Henao-Restrepo et al., 2017), we extended the chain-gamma-binomial model to exclude infections caused directly by the index case. This reduced the mean number of cases per cluster by ≈ 65%, lowering the power of fixed-size trials but leaving the qualitative pattern unchanged (Figure 5, bottom row).

Together, these results show that cluster-randomised ring-trials rapidly lose power as super-spreading intensifies, prompting us to explore whether individual-randomisation within rings could offer greater statistical efficiency.

### Super-Spreading Has Minimal Impact on Power of Individual-Randomised Designs

We next considered ring-trials in which each individual was randomised independently, regardless of the ring to which they belonged. Although individual-randomised trials are typically more powerful than cluster-randomised trials, it is important to quantify the extent of this advantage and how it depends on key factors. Several mechanisms influence the relative power of these designs: super-spreading increases intra-cluster correlation (ICC); *infection-blocking* vaccines modify the measured vaccine effectiveness through indirect effects; and indirect protection also reduces the total number of observed cases.

Figure 6 shows the power of a fixed-size individual-randomised ring-trial, with the same trial size and infection dynamics as in the cluster-randomised simulations (Figure 5). Individual-randomised designs consistently achieved greater power across all levels of super-spreading and vaccine efficacy, indicated by the larger blue regions in Figure 6. Moreover, the influence of super-spreading on power was minimal, with negligible impact until the most extreme levels (*k* < 0.1). This pattern of increased power and the lack of dependence on super-spreading was observed in all 4 metrics of power we evaluated (Figure S5 and S6).

**Figure 6:**
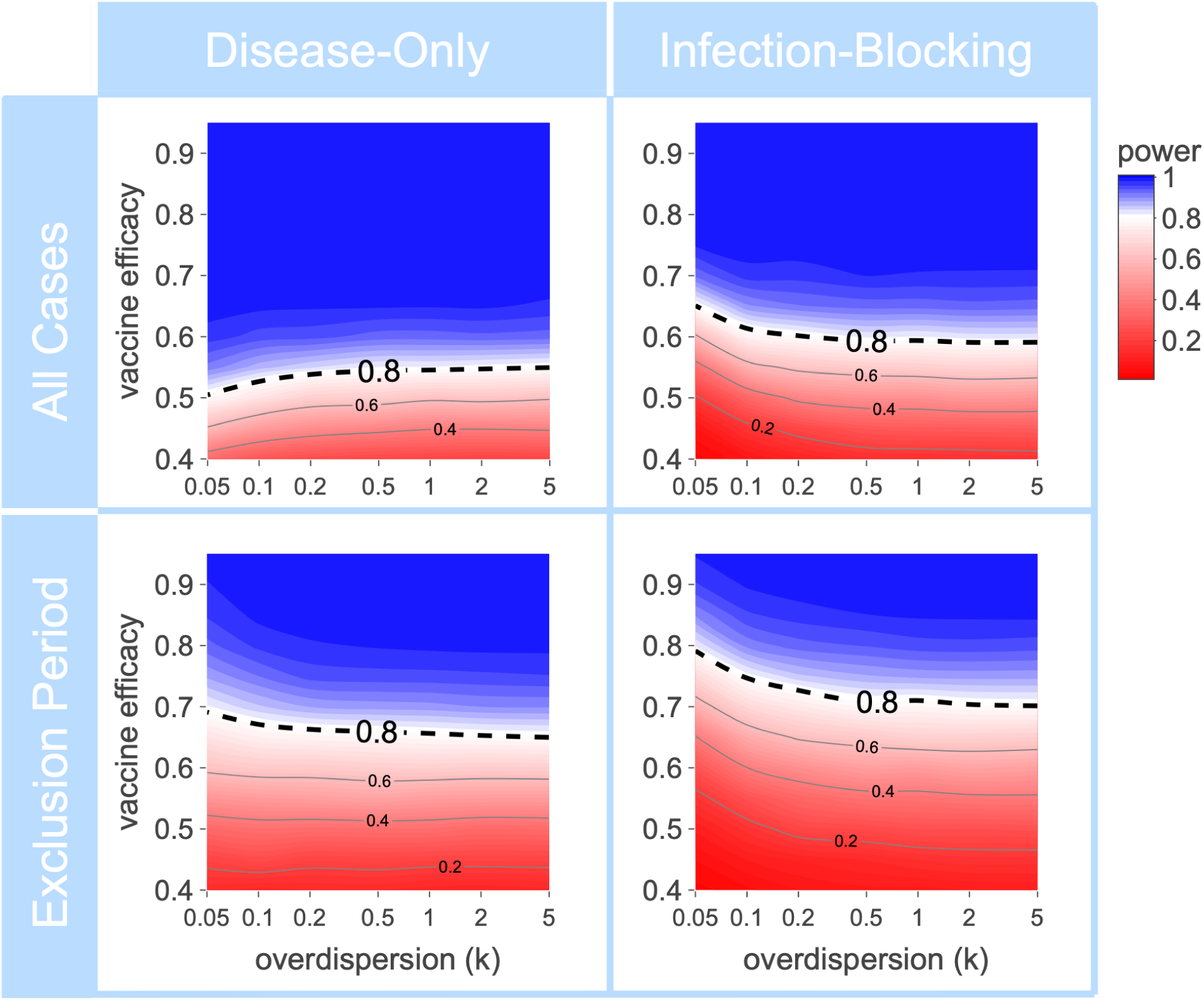
Super-spreading has a minimal effect on the power of individually-randomised trials. The probability of a fixed-size trial (200 rings; 30 per ring; 3.33% AR) demonstrating vaccine efficacy for different levels of super-spreading and vaccine efficacy. The individually-randomised trials are always more powerful than the cluster-randomised trials (compare the blue area with Figure 5). The power of the trial is approximately constant for different levels of super-spreading, with the exception of extreme levels (k<0.05), in contrast with cluster-randomised designs. The power of the trial is decreased if the vaccine is *infection-blocking* (right column) as opposed to a *disease-only* vaccine (left column), which is due to a reduction in the total number of within trial cases due to the indirect effect of the vaccine.

Interestingly, power decreased for *infection-blocking* vaccines compared with *disease-only* vaccines, the opposite pattern observed for cluster-randomised designs. In individual-randomised trials, indirect protection affects both trial arms equally, so the design measures only the direct vaccine efficacy. At the same time, the reduction in total cases due to indirect protection lowers power for a trial with a fixed number of rings. The trial power was also calculated using a Bayesian analysis of the chain-gamma-binomial models (see Methods) where the action of the vaccine (e.g. *disease-only* or *infection-blocking*) is part of the model. The results of the chain-gamma-binomial model and the beta-binomial model are almost identical (Figure S3)

## DISCUSSION

In this study, we analysed how super-spreading affects the statistical power of vaccine ring-trials, with a particular focus on the randomisation procedure. We found that high levels of super-spreading markedly reduce the power of cluster-randomised ring-trials, whereas individual-randomised ring-trials are largely unaffected. This difference arises because super-spreading increases the intra-cluster correlation (ICC), which diminishes the effective sample size and hence the power of cluster-randomised trials.

We also examined how vaccine mode of action influences trial performance. *Infection-blocking* vaccines increased the apparent power of cluster-randomised designs relative to *disease-only* vaccines due to indirect effects increasing the vaccine effectiveness, but the opposite effect occurred in individual-randomised designs. However, these differences were modest compared with the dominant impact of super-spreading, and individual-randomised ring-trials consistently achieved greater power across all scenarios. For pathogens with substantial super-spreading, such as a potential Nipah-X, cluster-randomised designs would only achieve adequate power if vaccine efficacy were nearly 100%.

The epidemic model used in this study was deliberately simple, but it explicitly incorporated super-spreading and different vaccine mechanisms. This structure enabled a comprehensive assessment of how these factors influence ring-trial design and allowed their relative effects on statistical power to be quantified. We therefore consider the conclusion, that cluster-randomised ring-trials are likely to fail for highly super-spreading pathogens unless vaccine efficacy is nearly perfect, to be generalisable beyond the specific case of Nipah-X. Although more detailed models, such as agent-based models, could provide additional insights, the extent of super-spreading represented within such frameworks would remain the dominant determinant of trial power. The parsimonious model presented here highlights this dependence clearly, revealing key mechanisms that might otherwise be obscured in more complex models.

An inherent challenge of the ring-trial design is that most cases occur within three to four weeks of identifying the index case. However, vaccines typically require at least a few days to confer protection and may provide little or no benefit to individuals already exposed before vaccination. To address this, ring-trials incorporate an exclusion period immediately after randomisation, during which cases are omitted from the efficacy analysis. This period is typically defined as a fixed cut-off of about one serial interval. In our model we represented this by excluding cases directly infected by the index case. The use of a fixed exclusion window is potentially problematic, since it must be pre-specified in the trial protocol and can strongly influence efficacy estimates. For example, in the *Ebola Ça Suffit!* trial, shortening the exclusion period from 10 to 7 days would have introduced two cases among vaccinated participants instead of none (Table S8; Henao-Restrepo et al., 2017), substantially lowering the apparent vaccine efficacy. An alternative is to weight cases by time since randomisation rather than using a fixed cut-off. This has been studied previously (Johnson et al., 2022), though without the complication of super-spreading. Evaluating how super-spreading influences these analytical methods represents an important direction for future research.

Ring-trial designs are often employed to evaluate vaccine efficacy when population infection incidence is low, with participants recruited based on their contact with confirmed index cases. In such trials, investigators must decide whether to randomise entire clusters or individuals within clusters. This decision requires balancing multiple factors, including trial power, logistical constraints and ethical considerations. Our analysis shows that for pathogens exhibiting super-spreading, individual-randomised ring-trials are substantially more powerful, whereas cluster-randomised designs achieve adequate power only when vaccine efficacy approaches 100%. These findings highlight the importance of accounting for super-spreading in trial design and suggest that individual-randomisation designs should be prioritised for evaluating vaccines against pathogens exhibiting super-spreading.

## METHODS

### Chain-Gamma-Binomial Model

The spread of infection in each ring is modelled using the chain gama-binomial model g (Fraser et al. 2011) extended to account for vaccination. Due to the high virulence of Nipah, we assume that each infection leads to a reported case with symptomatic disease. Each ring is modelled as a closed system of *N* initially-susceptible individuals with a single index-case who is the source of all first-generation infections. The chain model assumes that an infectious individual has the same probability of transmitting the infection to all susceptible members of the ring (*i.e.* the number of transmissions is binomially distributed). To model super-spreading, we assume that each infected individual has a different infectiousness which is independently drawn from a gamma distribution with shape parameter *k* and rate λ. The use of the gamma distribution for individual infectiousness is the same as the assumption used in the derivation of the negative-binomially distributed offspring distribution in branching-process models of super-spreading (Lloyd-Smith et al. 2005). The chain model then considers discrete generations of infections in the ring until the infection terminates. The effective reproduction number drops with each generation due to the depletion of susceptibles. Using the properties of the gamma distribution and the independence of the individual infectiousness, if there are *I_i_* individuals infected in the *i*^th^-generation, then the combined infection pressure on the remaining susceptibles in the ring will be gamma distributed with shape parameter *kI_i_* and rate λ. If there are *S_i_* susceptibles in the *i*^th^-generation, the distribution of infected individuals in the *i+1*^th^-generation is

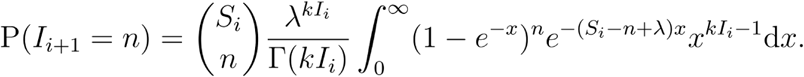

The final-size distribution is the total number of infected individuals across all generations (excluding the index case),

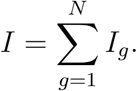

Using a Wald-identity, Ball (Ball, 1986) demonstrated that the final size distribution for a chain model satisfies

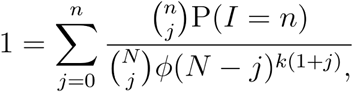

where *φ*(t) is the Laplace transform of the gamma distribution. This equation can be solved recursively to get the final-size distribution of rings where all individuals in the ring are either in the placebo or the vaccinated arm (i.e. cluster randomisation). In the model containing all generations, we simulate the number of end-points in each ring as independent draws from this distribution.

### Vaccine Model

Vaccines which are not 100% protective can be either *leaky* or *all-or-nothing* or a mix of both these effects. A *leaky* vaccine is where the vaccine reduces the risk of infection with each exposure, and an *all-or-nothing* vaccine is where a proportion of vaccinated individuals have complete protection from infection whilst others have zero protection. In this work we consider *leaky* vaccines.

*Infection-blocking* vaccines are modelled by reducing the mean infection pressure by a factor of *(1-v)* where *v* is the vaccine efficacy. This has the effect of reducing the total number of infections as well as symptomatic cases. *Disease-only* vaccines do not reduce the mean infection pressure; however, the probability that an infected vaccinated individual develops symptoms is *(1-v)*. We assume that following infection individuals develop sterilising immunity, so for *disease-only* vaccines the effect of *leaky* and *all-or-nothing* vaccines are identical. Thus, if there are *I* infections in a vaccinated ring, then the number of symptomatic cases *C* is

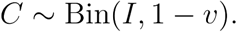

### Individually Randomised Rings

Next we consider individually randomised rings which contain *N_p_* individuals in the placebo arm and *N_v_* individuals in the vaccinated arm. Following the modelling assumptions of the chain gamma-binomial model, we assume all individuals within the ring interact with each other equally regardless of vaccination status. For a *disease-only* vaccine, the final-size for the total number of infected individuals is the same as that for a ring of size (*N_p_* +*N_v_*) with no vaccinations. Given that individuals in both arms are equally likely to be infected, then

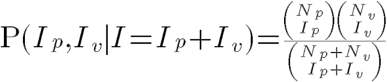

where *I_p_* and *I_v_* are the number of people infected in the placebo and vaccinated arms respectively. Since the vaccine is *disease-only*, the number of symptomatic vaccinated individuals *C_v_* is then

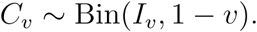

*Infection-blocking* vaccines are more complicated, with the vaccinated individuals within a ring feeling a lower infection pressure than the placebo individual in the same ring. Define *S_v,i_* and *S_p,i_*as the number of susceptibles in the vaccine and placebo arms in the *i*^th^-generation; and *I_v,i_*and *I_p,i_* as the number of new infections in the vaccine and placebo arms in the *i*^th^-generation. Then

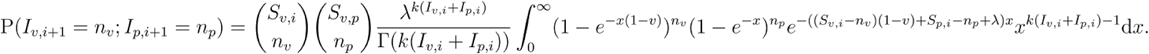

Using a Wald-identity, Ball (Ball, 1986) demonstrated that the final size distribution for a chain model with multiple groups satisfies

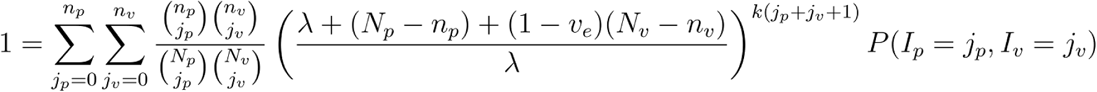

### Initial Exclusion Period

Index cases which are used to define rings will only be identified once they have developed symptoms and sought treatment. This will be followed by logistical delays in recruiting individuals to the trial, and delays in the vaccine conferring protection. Additionally, the incubation period between exposure and symptomatic disease is approximately 9.5 days (Arunkumar et al., 2020), therefore new cases in the rings identified immediately after randomisation are likely to have been infected prior to the vaccine taking effect. Therefore, new cases in the rings are excluded from the end-point if they develop symptoms in a window post-randomisation, for the *Ebola ca Suffit* trial this was set to 10 days, which is approximately one generation time. Whilst in principle all these features can be included in a model, the data is not available to calibrate them. Therefore we take a parsimonious approach and assume that the vaccine would not have had time to confer protection on individuals directly infected by the index case, and we exclude the first generation of cases from the trial end-point. For transmission from the first and subsequent generations we assume that the vaccine has taken effect and include new cases in the trial end-point.

### Bayesian Beta-Binomial Model

A super-spreading epidemic leads to the number of cases per ring being overdispersed compared to a binomial distribution. The simplest statistical model which can account for this is the beta-binomial model, where the infection pressure in each ring is beta distributed with parameters (*α*,*β*). If there are *N* individuals in a ring, then the probability of *n* infections in a ring is

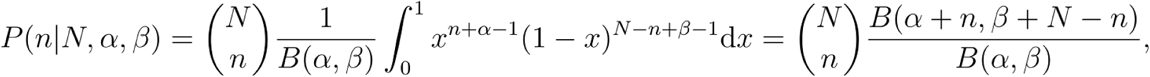

where *B*(*α*,*β*) is the beta function. For the cluster randomised design, the vaccine is assumed to reduce the infection pressure by a factor of *(1-v),* so the probability of n cases in a vaccinated cluster is

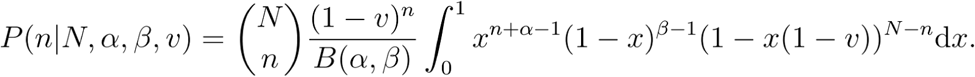

For the case of the individually randomised design, the vaccine reduces the infection pressure felt by the vaccinated individuals by a factor of *(1-v);* however, we have now both vaccinated and placebo individuals within each ring. If there are *N_v_*(*N_p_*) vaccinated (placebo) individuals in a ring, then the probability of *n_v_* (*n_p_*) infections amongst vaccinated (placebo) individuals in a ring is

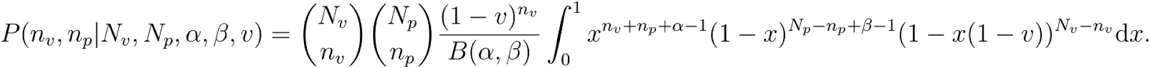

The (expected) attack rate and overdispersion can be expressed in terms of the parameters of the beta distribution

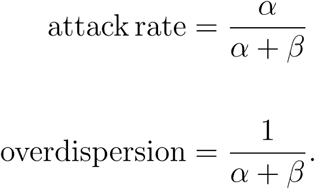

Weak priors were placed on attack rate, overdispersion and vaccine efficacy

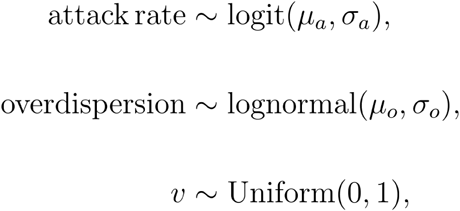

where *μ_0_ = -1, σ_o_= 2*, *σ_a_ = 1* and *μ_a_* is the mean attack rate across all the rings in the trial. The posterior distribution was sampled using the default MCMC algorithm in Stan (Stan Development Team, 2025). For each combination of vaccine efficacy and overdispersion parameter, 5,000 trials were simulated and results analysed using the Bayesian beta-binomial model. Trials proved vaccine efficacy if less than 5% of the posterior distribution of the vaccine efficacy was less than 30% and the power was the proportion of simulated trials which proved efficacy.

### Bayesian Chain-Gamma-Binomial Model

The likelihood function for the chain-gamma-binomal model and the extensions to include vaccines can be evaluated exactly by either matrix inversion or induction. Range priors were put on the vaccine efficacy, log(*kN/*λ) and log(*k*). The posterior distribution was sampled using the default MCMC algorithm in Stan (1000 posterior samples per simulation). Trials proved vaccine efficacy if less than 5% of the posterior distribution of the vaccine efficacy was less than 30% and the power was the proportion of simulated trials which proved efficacy.

### Software and Testing

All simulation and statistical models used in this paper are part of the Presto project, which is available as an R Package on Github (https://github.com/BDI-pathogens/presto). The model implementations have been extensively tested, including tests to show that Bayesian posterior distributions accurately cover the true simulation parameters over repeated model simulations.

## Data Availability

All data produced in the present study are available upon reasonable request to the authors

## SUPPLEMENTARY FIGURES

**Figure S1:**
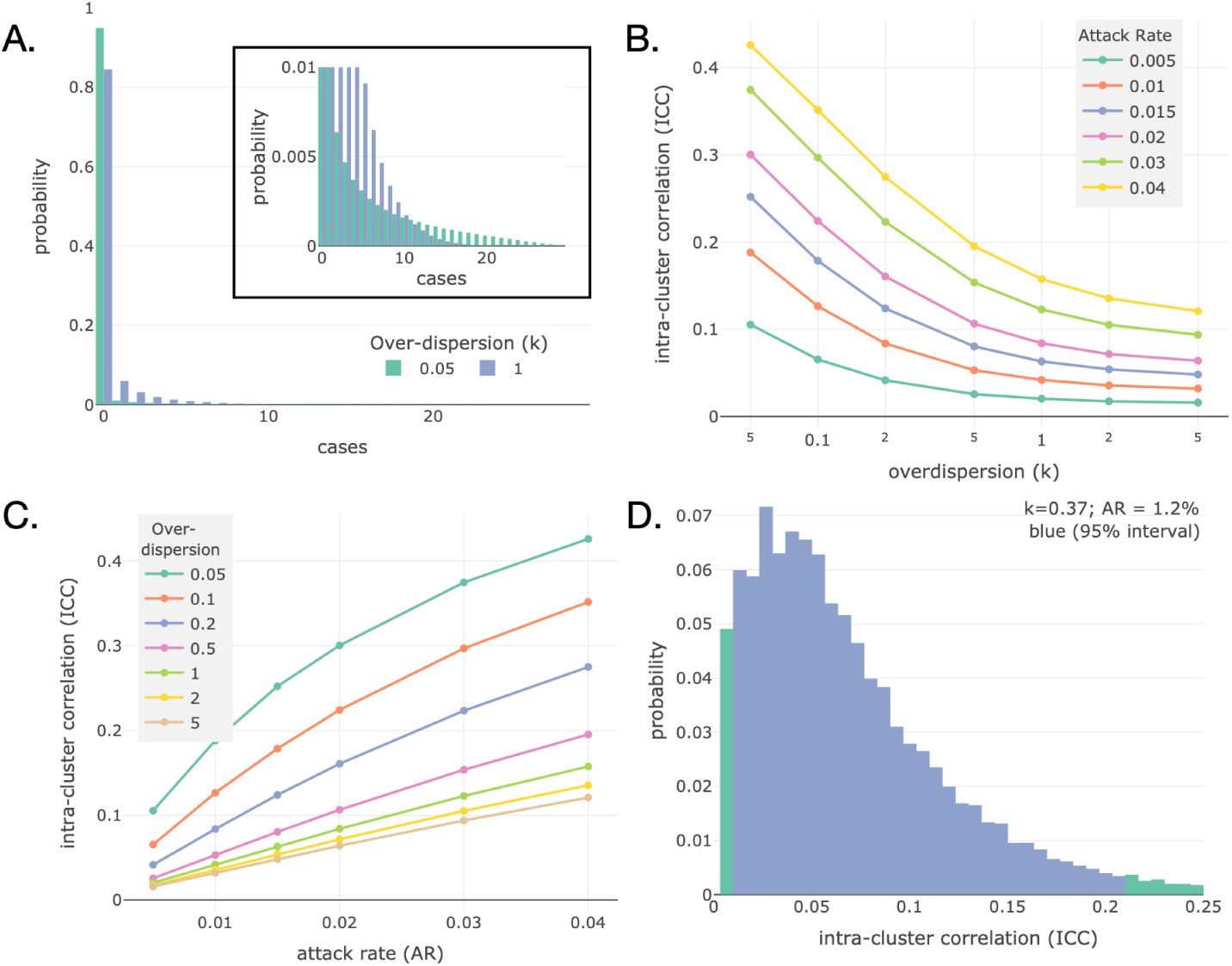
Super-spreading induces similar intra-cluster correlation when. **first-generation is omitted (compare with Figure 3).** A. The distribution of cases excluding the first generation in a ring of 30 individuals and an overall 3.33% AR. Excluding the first generation the attack rate is 1.16% which is 35% of the total cases. B. The ICC for clusters with different levels of super-spreading are slightly lower than when the first generation is included (Figure 3B). C. The ICC for clusters with different AR are slightly lower than when the first generation is included (Figure 3C). D. The distribution of the maximum-likelihood estimates of ICC for simulated trials of 47 rings each containing 65 individuals, using the AR from the *Ebola Ça Suffit!* trial and the estimate of k for Ebola. The 95% central interval spans 0%-20% demonstrating the large variation in ICC which can be observed in trials.

**Figure S2:**
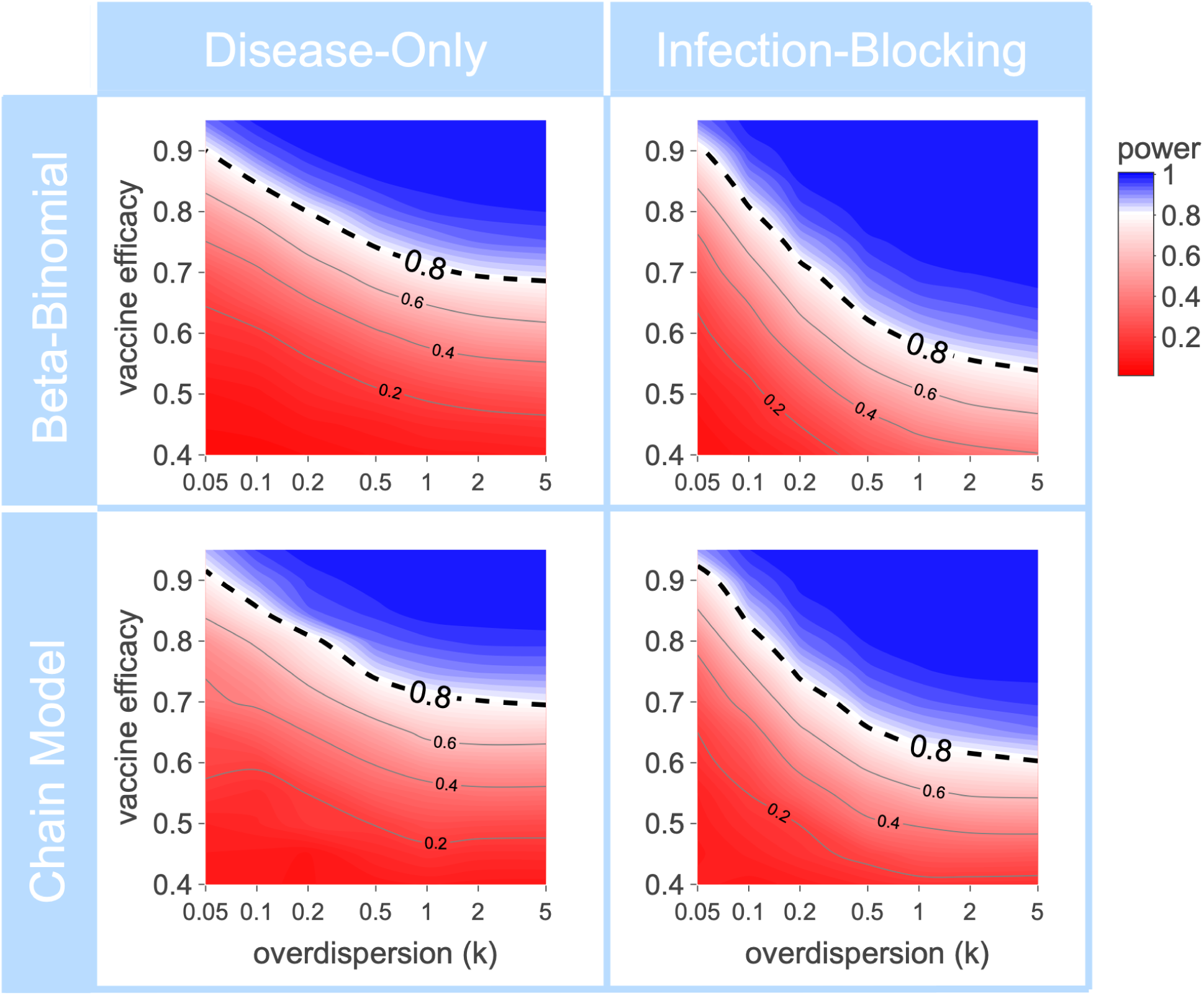
Comparison of beta-binomial and chain-gamma-binomial models for cluster randomised designs. The power of a cluster-randomised ring trial estimated using a beta-binomial (top row) and matched chain-gamma-binomial models (bottom row), for *disease-only* vaccines (left column) and *infection-blocking* vaccines (right column). The results are almost identical, with the exception that the beta-binomial model suggests the trial is more powerful to demonstrate VE with an *infection-blocking* vaccine. This is due to the beta-binomial model estimating the total vaccine effectiveness which is higher with *infection-blocking* vaccines due to indirect effects, whereas the chain-gamma-binomial model calculates the individual vaccine efficacy with the indirect effect implied by the model.

**Figure S3:**
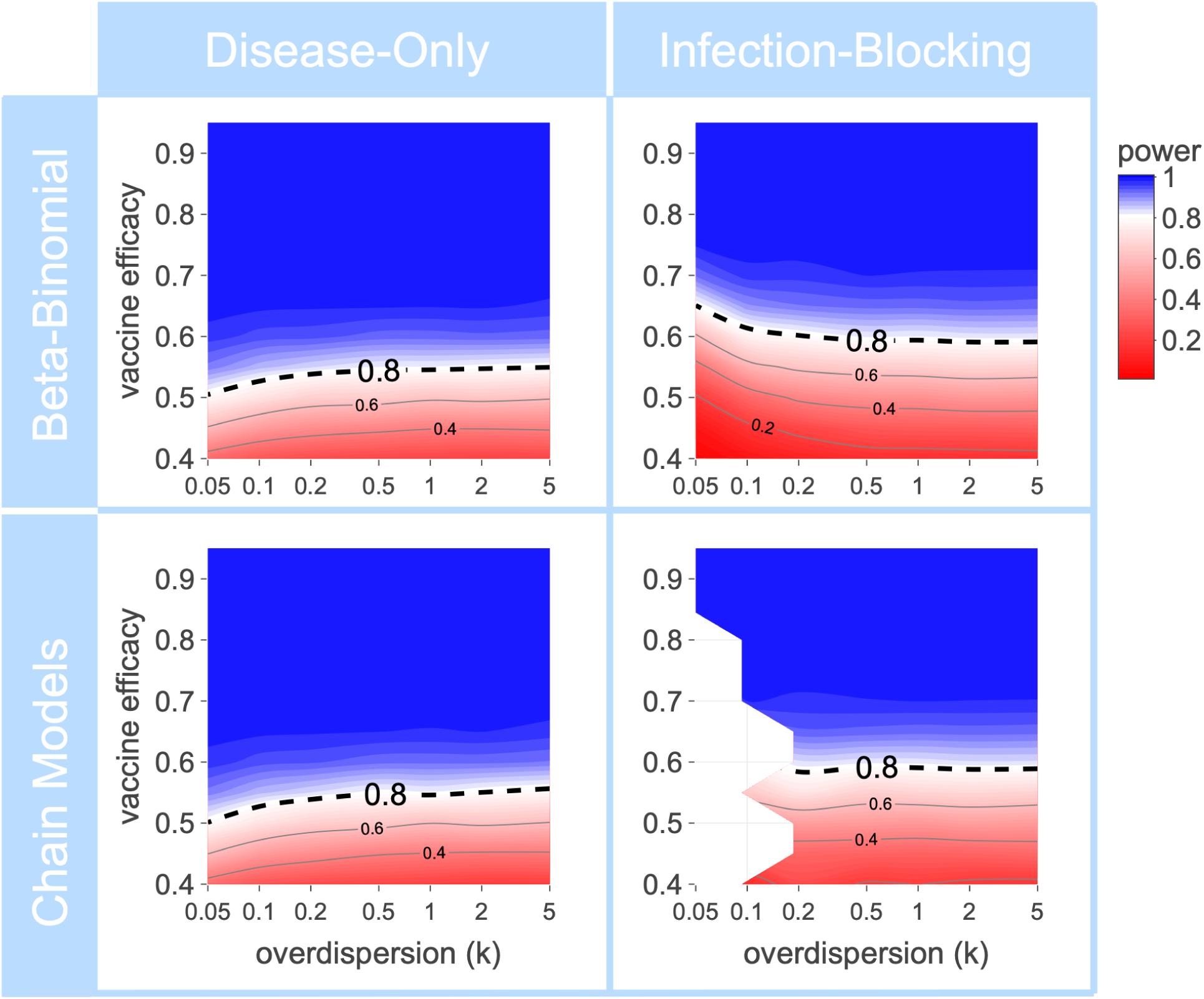
Comparison of beta-binomial and chain-gamma-binomial models for individual randomised designs. The power of an individually-randomised ring trial estimated using a beta-binomial (top row) and matched chain-gamma-binomial models (bottom row), for *disease-only* vaccines (left column) and *infection-blocking* vaccines (right column). The results of the 2 models are almost identical.

**Figure S4:**
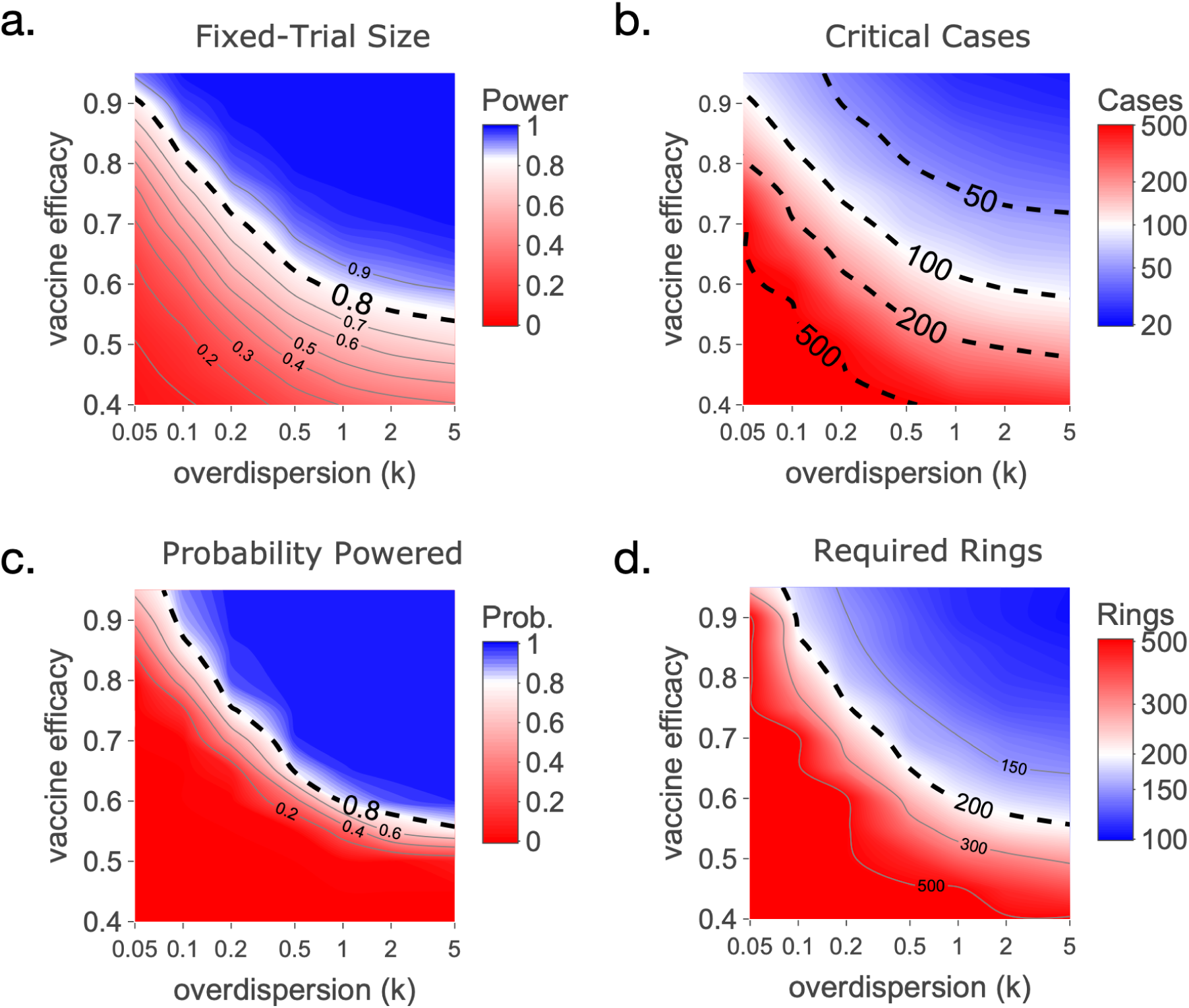
Measures of trial power. Different measures of trial power for an cluster-randomised ring-trial with an *infection-blocking* vaccine. a. the probability that a trial with a fixed number of rings (200 rings; 30 people per ring; AR 3.33%) is powered. b. the required number of cases across all rings such that the trial is 80% powered. c. the probability that a trial with a fixed number of rings accumulates the required cases. d. the required number of rings in the trial such that there is a 80% probability of accumulating the required cases. Each plot shows the trial power for different levels of super-spreading (low k mean high super-spreading) and vaccine efficacies, with the blue representing the region where trial is powered.

**Figure S5:**
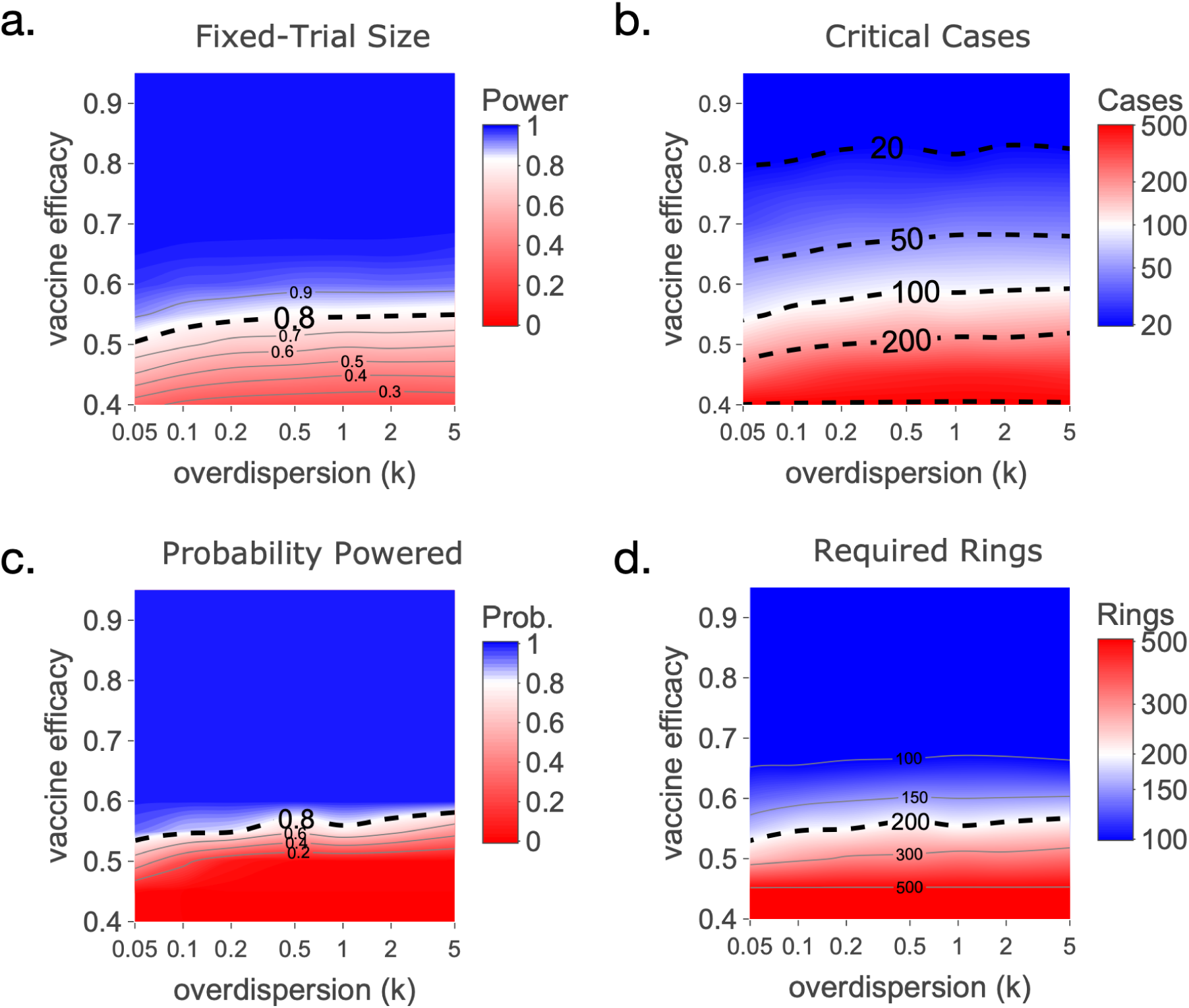
Measures of trial power. Different measures of trial power for an individually randomised ring-trial with a *disease-only* vaccine. a. the probability that a trial with a fixed number of rings (200 rings; 30 people per ring; AR 3.33%) is powered. b. the required number of cases across all rings such that the trial is 80% powered. c. the probability that a trial with a fixed number of rings accumulates the required cases. d. the required number of rings in the trial such that there is a 80% probability of accumulating the required cases. Each plot shows the trial power for different levels of super-spreading (low k mean high super-spreading) and vaccine efficacies, with the blue representing the region where trial is powered.

**Figure S6:**
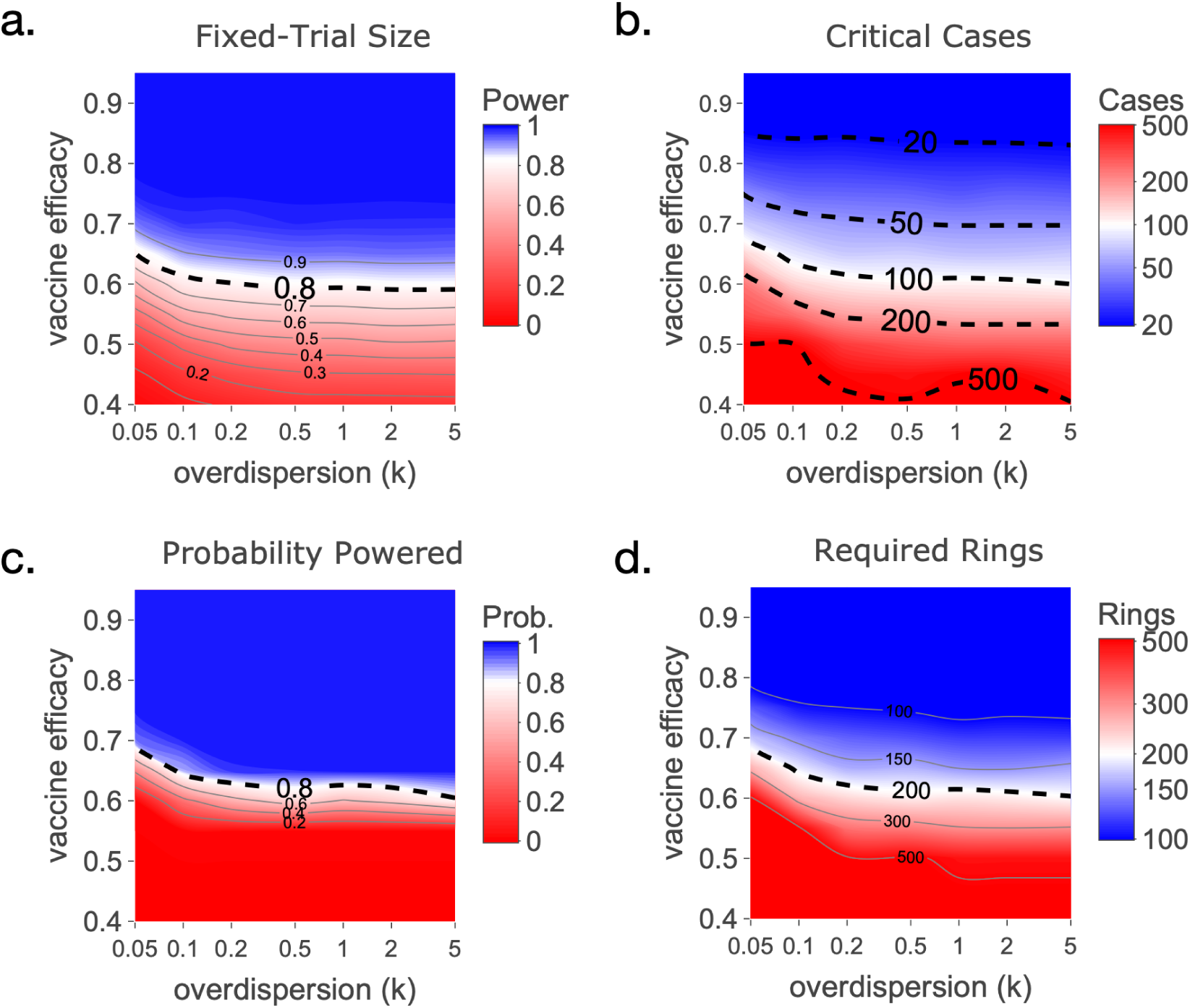
Measures of trial power. Different measures of trial power for an individually randomised ring-trial with an *infection-blocking* vaccine. a. the probability that a trial with a fixed number of rings (200 rings; 30 people per ring; AR 3.33%) is powered. b. the required number of cases across all rings such that the trial is 80% powered. c. the probability that a trial with a fixed number of rings accumulates the required cases. d. the required number of rings in the trial such that there is a 80% probability of accumulating the required cases. Each plot shows the trial power for different levels of super-spreading (low k mean high super-spreading) and vaccine efficacies, with the blue representing the region where trial is powered.

## REFERENCES

Antrobus RD, Coughlan L, Berthoud TK, Dicks MD, Hill AV, Lambe T, Gilbert SC. 2014. Clinical assessment of a novel recombinant simian adenovirus ChAdOx1 as a vectored vaccine expressing conserved Influenza A antigens. Mol Ther ;22(3):668–674. doi: 10.1038/mt.2013.284. Epub 2013 Dec 30. PMID: 24374965; PMCID: PMC3944330.

Arunkumar G, Chandni R, Mourya DT, Singh SK, Sadanandan R, Sudan P, Bhargava B; 2019. Nipah Investigators People and Health Study Group. Outbreak Investigation of Nipah Virus Disease in Kerala, India, 2018. J Infect Dis. ;219(12):1867–1878. doi: 10.1093/infdis/jiy612. PMID: 30364984.

Baden LR, et al. 2021. Efficacy and Safety of the mRNA-1273 SARS-CoV-2 Vaccine. N Engl J Med 2021;384:403–416 DOI: 10.1056/NEJMoa2035389

Ball F. A unified approach to the distribution of total size and total area under the trajectory of infectives in epidemic models. Advances in Applied Probability. 1986;18(2):289–310. doi:10.2307/1427301

Butzin-Dozier Z, Athni TS, Benjamin-Chung J. A Review of the Ring-Trial Design for Evaluating Ring Interventions for Infectious Diseases. Epidemiol Rev. 2022 Dec 21;44(1):29–54. doi: 10.1093/epirev/mxac003. PMID: 35593400; PMCID: PMC10362935.

CEPI 2023, CEPI and PATH strengthen partnership to accelerate development of vaccines against diseases with epidemic or pandemic potential. https://cepi.net/cepi-and-path-strengthen-partnership-accelerate-development-vaccines-against-diseases-epidemic-or

Chakraborty H, Hossain A. R package to estimate intracluster correlation coefficient with confidence interval for binary data. Comput Methods Programs Biomed. 2018 Mar;155:85–92. doi: 10.1016/j.cmpb.2017.10.023. Epub 2017 Oct 31. PMID: 29512507.

Coalition for Epidemic Preparedness Innovations (CEPI), 2022, Delivering pandemic vaccines in 100 days. https://static.cepi.net/downloads/2024-02/CEPI-100-Days-Report-Digital-Version_29-11-22.pdf

Colditz GA, Brewer TF, Berkey CS, Wilson ME, Burdick E, Fineberg HV, Mosteller F. Efficacy of BCG vaccine in the prevention of tuberculosis. Meta-analysis of the published literature. JAMA. 1994 Mar 2;271(9):698–702. PMID: 8309034.

Datoo, Mehreen; Mahamar, Almahamoudou et al. 2024. Safety and efficacy of malaria vaccine candidate R21/Matrix-M in African children: a multicentre, double-blind, randomised, phase 3 trial. The Lancet, Volume 403, Issue 10426, 533–544.

Dean, NA et al. 2019. Considerations for the design of vaccine efficacy trials during public health emergencies. Sci Transl Med .11(499): . doi:10.1126/scitranslmed.aat0360.

DeBuysscher BL, Scott D, Thomas T, Feldmann H, Prescott J. Peri-exposure protection against Nipah virus disease using a single-dose recombinant vesicular stomatitis virus-based vaccine. 2016. NPJ Vaccines. 2016;1:16002–. doi: 10.1038/npjvaccines.2016.2. Epub 2016 Jul 28. PMID: 28706736; PMCID: PMC5505655.

Fraser C, Cummings DA, Klinkenberg D, Burke DS, Ferguson NM. Influenza transmission in households during the 1918 pandemic. Am J Epidemiol. 2011 Sep 1;174(5):505–14. doi: 10.1093/aje/kwr122. Epub 2011 Jul 11. PMID: 21749971; PMCID: PMC3695637.

Gouglas D, Christodoulou M, Plotkin SA, Hatchett R. 2019. CEPI: Driving Progress Toward Epidemic Preparedness and Response. Epidemiol Rev. ;41(1):28–33. doi: 10.1093/epirev/mxz012. PMID: 31673694; PMCID: PMC7108492.

Henao-Restrepo, Ana Maria et al. 2017. Efficacy and effectiveness of an rVSV-vectored vaccine in preventing Ebola virus disease: final results from the Guinea ring vaccination, open-label, cluster-randomised trial (Ebola Ça Suffit!) The Lancet, Volume 389, Issue 10068, 505–518

Hodcroft EB, Wohlfender MS, Neher RA, Riou J, Althaus CL. Estimating Re and overdispersion in secondary cases from the size of identical sequence clusters of SARS-CoV-2. PLoS Comput Biol. 2025 Apr 15;21(4):e1012960. doi: 10.1371/journal.pcbi.1012960. PMID: 40233303; PMCID: PMC12040226.

Jacob ST, Crozier I, Fischer WA 2nd, Hewlett A, Kraft CS, Vega MA, Soka MJ, Wahl V, Griffiths A, Bollinger L, Kuhn JH. 2020. Ebola virus disease. Nat Rev Dis Primers. 6(1):13. doi: 10.1038/s41572-020-0147-3. PMID: 32080199; PMCID: PMC7223853.

Johnson R, Jackson C, Presanis A, Villar SS, De Angelis D. Quantifying Efficiency Gains of Innovative Designs of Two-Arm Vaccine Trials for COVID-19 Using an Epidemic Simulation Model. Stat Biopharm Res. 2022 Jan 2;14(1):33–41. doi: 10.1080/19466315.2021.1939774. Epub 2021 Jul 30. PMID: 35096276; PMCID: PMC7612285.

Killip S, Mahfoud Z, Pearce K. What is an intracluster correlation coefficient? Crucial concepts for primary care researchers. Ann Fam Med. 2004 May-Jun;2(3):204–8. doi: 10.1370/afm.141. PMID: 15209195; PMCID: PMC1466680.

Longini IM Jr, Halloran ME, Haber M, Chen RT. 1993. Measuring vaccine efficacy from epidemics of acute infectious agents. Stat Med. ;12(3-4):249–63. doi: 10.1002/sim.4780120309. PMID: 8456210

Lau MS, Dalziel BD, Funk S, McClelland A, Tiffany A, Riley S, Metcalf CJ, Grenfell BT. Spatial and temporal dynamics of superspreading events in the 2014-2015 West Africa Ebola epidemic. Proc Natl Acad Sci U S A. 2017 Feb 28;114(9):2337–2342. doi: 10.1073/pnas.1614595114. Epub 2017 Feb 13. PMID: 28193880; PMCID: PMC5338479.

Lloyd-Smith JO, Schreiber SJ, Kopp PE, Getz WM. Superspreading and the effect of individual variation on disease emergence. Nature. 2005 Nov 17;438(7066):355–9. doi: 10.1038/nature04153. PMID: 16292310; PMCID: PMC7094981.

Maniscalco, D, et al. 2024. Adaptive behavior in response to the 2022 mpox epidemic in the Paris region. medRxiv 2024.10.25.24315987; doi: 10.1101/2024.10.25.24315987

Nikolay B, Salje H, Hossain MJ, Khan AKMD, Sazzad HMS, Rahman M, Daszak P, Ströher U, Pulliam JRC, Kilpatrick AM, Nichol ST, Klena JD, Sultana S, Afroj S, Luby SP, Cauchemez S, Gurley ES. 2019. Transmission of Nipah Virus - 14 Years of Investigations in Bangladesh. N Engl J Med;380(19):1804–1814. doi: 10.1056/NEJMoa1805376. PMID: 31067370; PMCID: PMC6547369.

Nikolay B, Salje H, Khan AKMD, Sazzad HMS, Satter SM, Rahman M, Doan S, Knust B, Flora MS, Luby SP, Cauchemez S, Gurley ES. A Framework to Monitor Changes in Transmission and Epidemiology of Emerging Pathogens: Lessons From Nipah Virus. J Infect Dis. 2020 May 11;221(Suppl 4):S363–S369. doi: 10.1093/infdis/jiaa074. PMID: 32392322; PMCID: PMC7213557.

Nikolay B, Ribeiro Dos Santos G, Lipsitch M, Rahman M, Luby SP, Salje H, Gurley ES, Cauchemez S. 2021. Assessing the feasibility of Nipah vaccine efficacy trials based on previous outbreaks in Bangladesh. Vaccine. 2021 Sep 15;39(39):5600–5606. doi: 10.1016/j.vaccine.2021.08.027. Epub 2021 Aug 20. PMID: 34426025.

Osterholm MT, Kelley NS, Sommer A, Belongia EA. Efficacy and effectiveness of influenza vaccines: a systematic review and meta-analysis. Lancet Infect Dis. 2012 Jan;12(1):36–44. doi: 10.1016/S1473-3099(11)70295-X. Epub 2011 Oct 25. Erratum in: Lancet Infect Dis. 2012 Sep;12(9):655. PMID: 22032844.

Pardi, N., Hogan, M., Porter, F. et al. 2018. mRNA vaccines — a new era in vaccinology. Nat Rev Drug Discov 17, 261–279.. 10.1038/nrd.2017.243

Pischel L, Martini BA, Yu N, Cacesse D, Tracy M, Kharbanda K, Ahmed N, Patel KM, Grimshaw AA, Malik AA, Goshua G, Omer SB. Vaccine effectiveness of 3rd generation mpox vaccines against mpox and disease severity: A systematic review and meta-analysis. Vaccine. 2024 Nov 14;42(25):126053. doi: 10.1016/j.vaccine.2024.06.021. Epub 2024 Jun 21. PMID: 38906763.

Rodrigue V, Gravagna K, Yao J, Nafade V, Basta NE. 2024. Current progress towards prevention of Nipah and Hendra disease in humans: A scoping review of vaccine and monoclonal antibody candidates being evaluated in clinical trials. Trop Med Int Health. 2024 May;29(5):354–364. doi: 10.1111/tmi.13979. Epub 2024 Feb 28. PMID: 38415314.

Stan Development Team, 2025. RStan: the R interface to Stan. R package version 2.36.0.9000. https://mc-stan.org/.

van Doremalen N, Avanzato VA, Goldin K, Feldmann F, Schulz JE, Haddock E, Okumura A, Lovaglio J, Hanley PW, Cordova K, Saturday G, de Wit E, Lambe T, Gilbert SC, Munster VJ. ChAdOx1 NiV vaccination protects against lethal Nipah Bangladesh virus infection in African green monkeys. NPJ Vaccines. 2022 Dec 21;7(1):171. doi: 10.1038/s41541-022-00592-9. PMID: 36543806; PMCID: PMC9768398.

Voysey, MerrynAban, Marites, et al. 2021. Safety and efficacy of the ChAdOx1 nCoV-19 vaccine (AZD1222) against SARS-CoV-2: an interim analysis of four randomised controlled trials in Brazil, South Africa, and the UK. The Lancet, Volume 397, Issue 10269, 99–111

WHO, 2024. CORE Protocol - A phase 1/2/3 study to evaluate the safety, tolerability, immunogenicity, and efficacy of vaccine candidates against (Filoviruses) virus disease in healthy individuals at risk of (Filovirus) virus disease. https://www.who.int/publications/m/item/core-protocol-a-phase-1-2-3-study-to-evaluate-the-safety-tolerability-immunogenicity-and-efficacy-of-vaccine-candidates-against-filoviruses-disease-in-healthy-individuals-at-risk-of-filovirus-disease

Yadav, P.D., Baid, K., Patil, D.Y. et al. 2025 A One Health approach to understanding and managing Nipah virus outbreaks. Nat Microbiol 10, 1272–1281. 10.1038/s41564-025-02020-9

Ypma RJ, Altes HK, van Soolingen D, Wallinga J, van Ballegooijen WM. A sign of superspreading in tuberculosis: highly skewed distribution of genotypic cluster sizes. Epidemiology. 2013 May;24(3):395–400. doi: 10.1097/EDE.0b013e3182878e19. PMID: 23446314.

